# Behavioral Clusters and Lesion Distributions in Ischemic Stroke, based on NIHSS Similarity Network

**DOI:** 10.1101/2023.11.08.23297808

**Authors:** Louis Fabrice Tshimanga, Andrea Zanola, Silvia Facchini, Antonio Luigi Bisogno, Lorenzo Pini, Manfredo Atzori, Maurizio Corbetta

## Abstract

**Purpose:** Stroke is a leading cause of death and disability. The subsequent dysfunctions relate to brain lesion location. The complex relationship between behavioral symptoms and lesion location is essential for care and rehabilitation, and useful to understand the healthy brain. Such complexity eludes linear methods. Differently from previous works, this study clusters patients by recurrent symptoms profiles, then retrieves their neural correlates.

**Methods:** Prevalence, severity and associations of symptoms affecting patients are condensed in a similarity measure between their profiles. The computation specifically adheres to the ordinal nature of NIHSS data. A network model links patients with strength proportional to the behavioral similarity. The network nodes are then classified through a novel spectral clustering variation. Lesions from each cluster’s patients are used in a voxel-wise analysis to statically validate the findings.

**Results:** The behavioral clusters express symptoms co-occurring in accordance with the literature on behavioral deficits covariance. Moreover, they show coherent groups of brain lesions in the associated CT and MRI scans. The anatomical position of the significant voxels found are aligned with the literature. Even when lesion density maps overlap, the significant voxels are well separated.

**Conclusion:** The presented workflow offers concrete, clinically useful correlations between patients profiles, individual patients-to-group associations, in-group co-occurrences of symptoms, and brain lesion correlates. The underlying mathematical assumptions, and the unsupervised machine learning techniques offer statistically robust results and are being further applied to other multimodal biomedical data.

## 1 Introduction

Stroke is the second leading cause of death and the third leading cause of disability worldwide [1] [2]. It occurs when there is an interruption in oxygenation to the brain, leading to cell death. In ischemic stroke, blood clots interrupt blood flow, whereas in hemorrhagic stroke, vessels break and blood diverges from its intended path. The subsequent damage to the brain of survivors can result in acute and chronic dysfunctions across sensory, motor, cognitive and behavioural domains. For instance, focal cortical lesions in highly specialized brain regions can affect narrow sets of functions, as in Broca’s and Wernicke’s aphasias. More often, subcortical strokes damage both grey and white matter, disrupting connections across the brain beyond the lesion location. These phenomena shed light and spark debates on the functional organization of the brain.

To characterize and quantify behavioral symptoms, clinicians have developed several tests that score performance in multiple domains, such as the Montreal Cognitive Assessment (MoCA) [3], the Oxford Cognitive Screen (OCS) [4] and the National Institutes of Health Stroke Scale (NIHSS) [5] [6]. The NIHSS is a 15 item scale that rates the severity of sensory, motor, attention and language deficits in stroke survivors. A score of 0 signifies the absence of deficit and healthy response, while maximum damage values range from 2 to 4, depending on the item. For example, the motor arm test involves evaluating the ability to hold the limb against gravity; 0 is given when there is no drift (limb hold for full 10 seconds at 45 or 90 degrees), while 4 is given when the patient is unable to move the limb; each limb has its respective NIHSS item. The NIHSS test can be quickly administered at admission, during the acute phase, at the neuropsychological evaluation, and later on during recovery. Many studies have focused on the sum of all item scores, while analyses of the separate scores have often characterized only inter-item correlations at the population level.

Such analyses have repeatedly identified a low-dimensional structure of the behavioral deficits’ covariance, with 2 up to 5 [7] [8] [9] [10] [11] components explaining the larger share of variance. These factors of variability separately correlate with lateral motor impairments, language, memory, and attention. Regularized regression models, statistical tests and machine learning techniques have been developed to relate behavioural scores to brain scans.

Despite several studies have analysed the relationship between lesion locations and behavioural symptoms, some delimiting aspects should be highlighted. First, many studies rely on implicit mathematical assumptions that might distort or discard information. In particular, although the use of rank correlations and tests is settled [12] [13], ordinal scores as found in NIHSS are usually treated as counts or continuous intervals [14] [15]. Alternatively, ordinal scales are reduced to dichotomous variables, e.g. *{*0, 1*}* to denote absence or presence of deficits [16] [17], or the cumulative sum of scores is thresholded to separate into classes of milder impairment and classes of worse prognosis [15] [18].

Second, the relationships among item scores [7] [8] and with lesion volumes and locations are usually modeled with linear methods, both for ordinal [19] and continuous scores [20].

Third, the scope is usually to study global, population-level associations between single deficits, rather than conditional associations and multivariate distributions of symptoms, i.e. syndromes. The former perspective, while insightful especially when developing clinical tests, is not concerned with individual-level phenotypization, nor sufficient for prediction of groups of deficits. In this regard, it is also notable that predicting deficits from lesions aligns with the aetiological process, but reverses the chronological order of assessment and availability typical of the clinical setting.

This study complements previous perspectives by introducing several innovative approaches. First, a network model of pairwise similarities among subjects is presented, with a conservative distance measure with regards to the ordinal nature of data, better suited than traditional alternatives such as Euclidean or Manhattan/Hamming distances between score vectors. Second, cluster-specific lesion maps arise naturally from clustering analysis by averaging lesion masks from patients in each cluster, rather than modeling correlations between latent variables derived from dimensionality reduction of behavioral and image variables. Third, a statistical evaluation and refinement of lesion mapping is conducted to identify lesions that distinguish behaviorally different groups. This approach assists in understanding aetiology and allows to align the complete profile of a single patient to a more general phenotype or syndrome, rather than associating a single symptom across the whole population to other single symptoms.

As mentioned in Yang et. al (2023) [21], clustering is one of the most useful methods for analyzing patient similarities for precision medicine. It groups patients into clinically meaningful subsets, which can be used for a variety of tasks, such as personalized therapies and policy making. Kim et. al (2022) [22] stress that finding similar clusters among stroke patients can be helpful from a medical perspective, as it may lead to the discovery of new patterns and more effective ways to manage stroke.

Overall, clustering is an effective method to associate many patients with each other, and to a few behavioral syndromes; to associate behavioral syndromes to specific lesion locations.

## 2 Materials and Methods

### 2.1 Data

The NIHSS data consist of 15 items describing the health state, abilities, and cognitive functions of patients. Some items are based on exterior observations that assess damage in specific areas of the brain, while other items test patients with more complex tasks, possibly involving disparate brain regions. NIHSS scores here refer to the acute phase. Statistics of NIHSS items are computed on first-stroke ischemic patients from Padua University Hospital and from Washington University in St. Louis data sets (total *n* = 308), while patients clustering is performed on the subset with both NIHSS> 0 (*n* = 248) and available brain scans (*n* = 172). While the latter enables symptoms-tolesions mapping that would otherwise be impossible, the initial selection implies that a total NIHSS score of 0 results from lesions that do not affect function in a measurable way. The average time between the stroke event and the NIHSS assessment for the *n* = 189 ischemic subjects in Padua is M = 3, SD = 3 days, while for the *n* = 119 ischemic subjects in St. Louis is M = 13, SD = 5 days. The Level Of Consciousness (LOC)-Vigilance item is removed, since the only subject not scoring 0 is filtered out, to exclude incomplete results from unresponsive patients.

Imaging data come from CT and MRI scans, co-registered in MNI152 standard space [23] at 1mm^3^ resolution, with lesions segmented by professionals. Combining cohorts and filtering, the resulting data set comprises 172 patients, whose ages ranged from 21 to 94 years (M=64, SD=15 years). A summarizing scheme of data selection is shown in Figure 1.

**Fig. 1.**
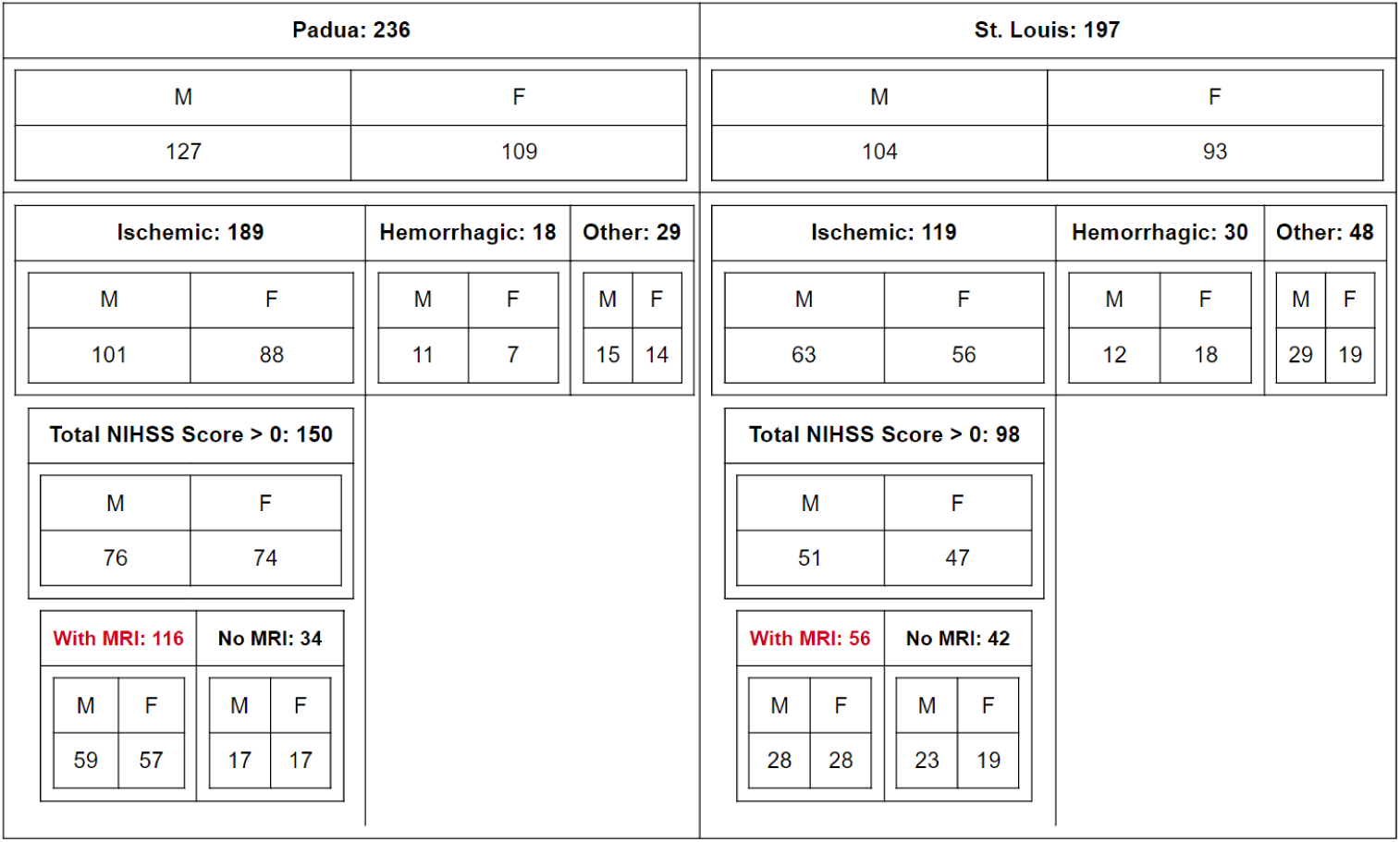
Patient selection workflow. Red marks patients taken into account for clustering. Abbreviations: M: male, F: female.

### 2.2 Correlations and Distances

Denoting *P* the set of *n* patients, and *X_n_*_=308_*_,m_*_=14_ the matrix of NIHSS observations considered, the relationship between NIHSS items across the population is explored with statistics suitable for ordinal data. Co-occurrences of deficits are counted irrespective of severity: for each couple of items, the number of patients exhibiting both deficits is counted. Then Spearman’s rank correlation coefficient among items is computed, to measure positive and negative associations. These are all operations of type *XT X*_(_*_m_*_=14_*_,m_*_=14)_.

Subsetting *P* to the *n* patients with NIHSS> 0 and available brain scans, *X_n_*_=172_*_,m_*_=14_ is the matrix of NIHSS observations for subsequent analyses. The General Distance Measure [24] [25] (GDM) is the proximity measure quantifying how different two patients are, based on their profile composed of 14 NIHSS scores. The GDM is defined taking inspiration from Kendall’s General Correlation Coefficient, in order to process continuous as well as ordinal values. It is here corrected and simplified in notation as:

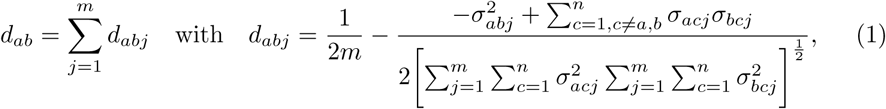

and

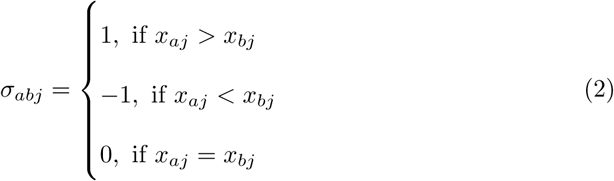

with

*a, b, c* = 1, 2, …, *n*: indexes of the *n* subjects (stroke patients)
*j* = 1, 2, …, *m*: index of the m variables (NIHSS items)
*x_aj_*: value of the variable j for subject a (NIHSS score)
σ*_abj_*: sign of the difference between patient a and patient b for item j d*_abj_*: distance between subjects a and b, on item j.
*d_ab_*: distance between subjects a and b.

Note that the GDM and its complement, the General Similarity Measure (GSM, of value *s_ab_* := 1 *− d_ab_*), depend on the whole cohort observations *X* for each pairwise computation. These are operations of type 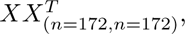 thus computing similarities between patients can be seen as the dual problem with respect to the problem of computing correlations between variables. The matrix of all pairwise relationships is symmetric, describing an undirected weighted graph: it is hence called the adjacency matrix *W*_(_*_n_*_=172_*_,n_*_=172)_ for the graph *G*(*P, W*), with *n* nodes referring to the patients and *n*(*n* − 1)/2 weighted edges referring to the GSM of all pairs of patients. For simplicity, *W* will hereon denote the matrix or the network, depending on context.

### 2.3 Repeated Spectral Clustering

Repeated Spectral Clustering (RSC) is a novel consensus clustering approach, defined in this paper based on spectral clustering, and aimed at dealing with the variability of results that stems from random initializations. Spectral clustering is the name for techniques based on the spectrum of the adjacency matrix’s (or graph’s) Laplacian [26]. Further arguments for using spectral clustering instead of other common techniques, such as hierarchical clustering and *k*-means, can be found in supplementary material Appendix B. Spectral clustering consists in two steps: spectral embedding, and clustering in the embedding space. The spectral embedding step yields Euclidean coordinates for the network nodes, in the space defined by the eigenvectors of the graph Laplacian. The actual clustering step can be any algorithm working in Euclidean spaces, typically *k*-means in embedding space [27], as in the present case. RSC is also a consensus clustering technique, with two distinct phases of spectral clustering. In the first phase there is evidence accumulation: the results of *N* different spectral clustering runs on *W* are recorded in the (*n × n*) consensus matrix *C*. In the second phase, *C* itself undergoes spectral clustering [28]. RSC is defined by the use of *L_sym_* spectral clustering [27] for both phases, whereas other consensus clustering methods change algorithm between the two phases, or change algorithm and/or data subset in the *N* runs of the first phase. While the entries of *W* measure the similarities between any two subjects, the corresponding entries of *C* count how many times those any two subjects are clustered together over the *N* runs of the evidence accumulation phase [29]. The more times two subjects are found in the same cluster, the more evidence those subjects are actually similar and co-members of the “real” cluster. Matrix *C* too induces a weighted undirected graph and its spectral properties and visual aspect highlight how the subject nodes are better separable than they are in *W*. Defined in this way, RSC unifies the ability to find clusters in spectral embedding with the robustness of results regarding centroids initialization, thanks to the evidence accumulation stage. As with other algorithms akin to *k*-means, the number of clusters *k* is set a priori or heuristically: here, the methodology relies on the spectral properties of *C* as a function of *k*. Overall, RSC can be interpreted as a way to extract the clustering signal *C* in the noisy *W*; averaging multiple measurements. The modules of RSC are described in Algorithm 1, Algorithm 2, and Algorithm 3 for spectral embedding, clustering and the whole RSC respectively. For further details refer to Appendix C.

#### Algorithm 1 spectral embedding

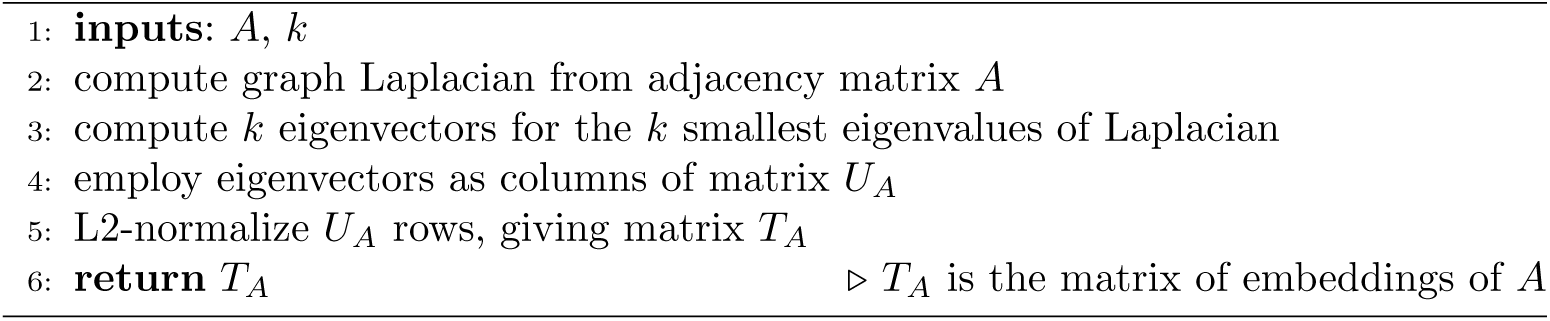

#### Algorithm 2 clustering

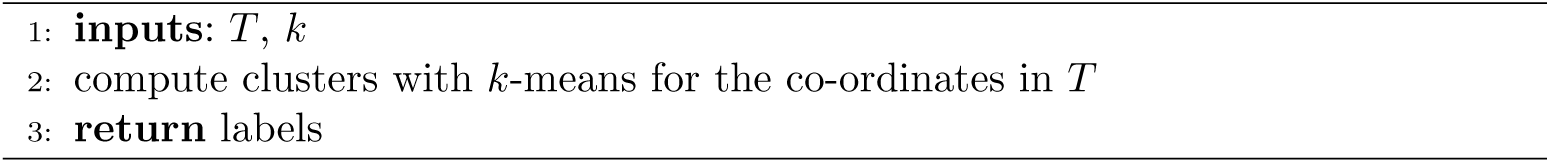

#### Algorithm 3 Repeated Spectral Clustering

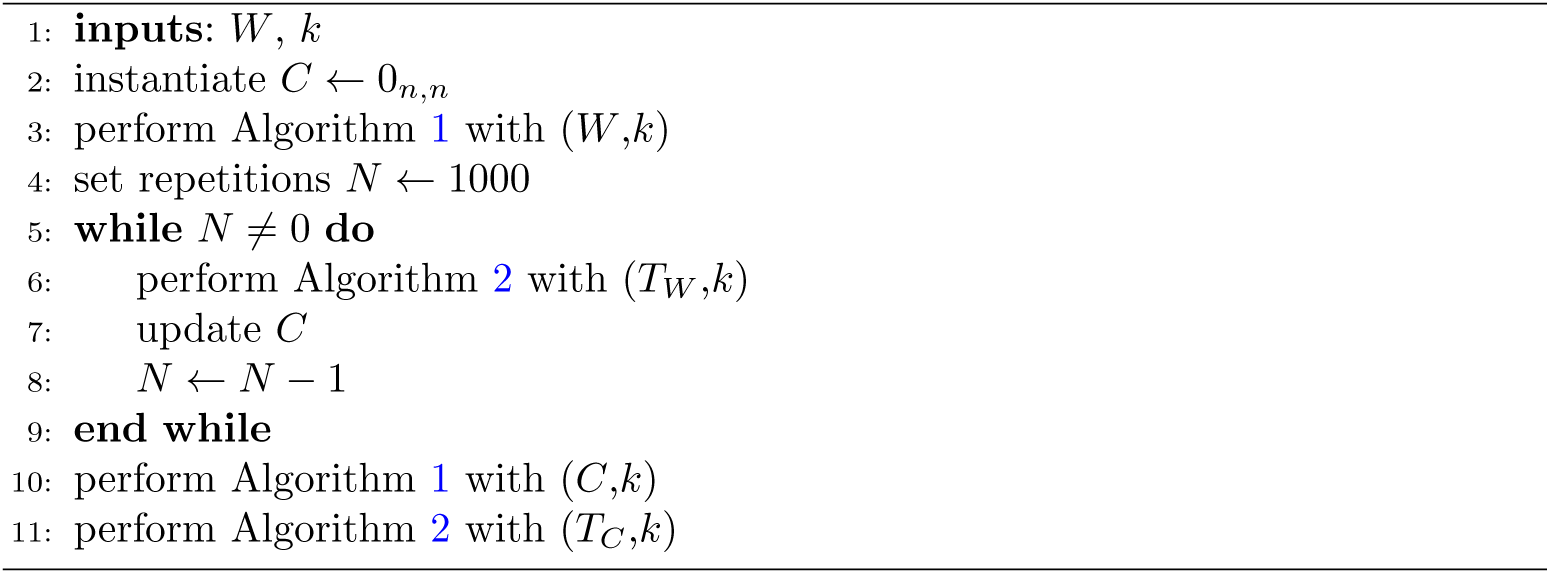

### 2.4 Voxel-wise analysis

The application of RSC to the affinity matrix obtained from the GDM pairwise similarities, leads to the isolation of clusters. In order to evaluate how a cluster is different from the others, lesion density maps can be visually compared in Figure 3, Panel **B**. Each map is obtained averaging the binary lesion masks in the respective cluster: thus, each voxel intensity is the [0-1] normalized frequency with which that voxel is lesioned in the cluster considered; equivalently, it is the normalized frequency of patients in that cluster that have a lesion in that location. To quantitatively test how significant are differences between clusters, we further perform a voxel-wise analysis. Since the objective of the clustering algorithm is to separate clusters in the space related to NIHSS items, statistical test on the separation in these same variables would lead to inflated p-values. For this reason, the statistical analysis focuses on voxel that are lesioned with significantly different frequencies across clusters.

The analysis conducted is composed by the following steps:

1. The comparisons are done in a one vs all-but-one way; the null-hypothesis *H*_0_ affirms that the lesion distribution in one cluster is not different from the lesion distribution of all the remaining patients.
2. From the 182 *×*218 *×*182 available voxels in MNI-152 template, we select those that are lesioned at least 8 times, which corresponds to the 5% of the patients. This is both for a statistical reason, since these voxels carry a greater effect size, as well as a computational reason, since this selection considers only 241163 voxels.
3. The test performed is the difference between proportions with pooled variance [30]:

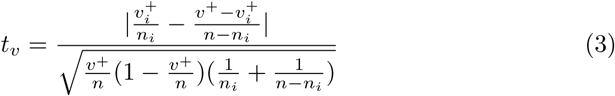 where:
  *n_i_*: number of patients in cluster i.
  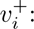 number of times voxel v, is lesioned (+) considering patients in cluster i.
  *n*: number of patients (172).
  *v*^+^: number of times voxel v, is lesioned (+) considering all the patients.
4. A permutational approach is used, where patients’ cluster assignment is randomly shuffled; for each selected voxel the statistic is computed 5000 times. In this way the distribution of the statistic *t_v_* can be built under *H*_0_, where there is random assignment of patients lesions to clusters.
5. Due to the large number of hypotheses under test (1 for each voxel), a multiple-comparison correction is needed. Both Bonferroni and Holm’s [31] methods are largely conservative, so a multistep-maxT (Westfall & Young [32]) procedure is used instead. This allows a Family Wise Error Rate (FWER) control at α = 0.01.

Since RSC is performed with *k*=5, the presented methodology is applied five times. These other five comparisons are not taken into account by the max-T algorithm; consequently, a Bonferroni correction is applied. Therefore, the results are said to be overall corrected, with FWER at *α* = 0.05.

### 2.5 Open-source software tools

Filtering data, statistical analyses, clustering, lesion frequency map computation and visualization are all performed with the Python 3 programming language, and its libraries numpy, pandas, scipy, scikit-learn, nibabel, and nilearn. Voxel-wise analysis are conducted with the R programming language, and its libraries foreach, doFuture, utils, scales, reticulate. The multi-step maxT correction is done using the code available at https://github.com/livioivil/r41sqrt10. In order to allow the usage of the presented analytic workflow by the scientific community on different datasets, code will be available at https://github.com/MedMaxLab/nihss_clustering; data will be available upon reasonable request to the authors.

## 3 Results

### 3.1 Correlations

The first set of results illustrates the co-occurrence of deficits, i.e. the contemporary presence regardless of severity scored, and the rank correlations of deficits across subjects, accounting for their severity (scores ranging from 0-2, to 0-4 for items in the NIHSS). Figure 2, Panel **A** shows the co-occurrence deficit matrix.

**Fig. 2.**
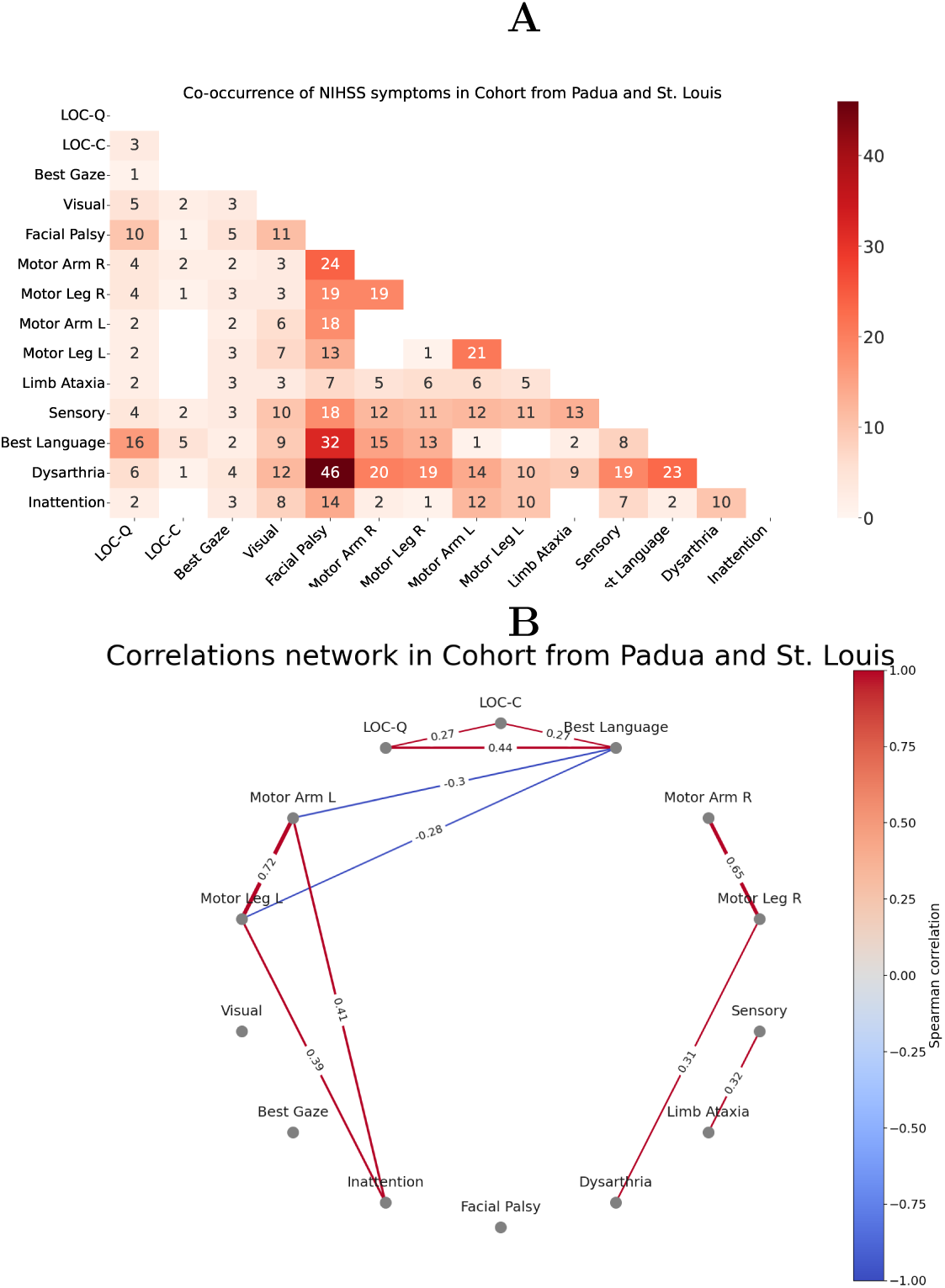
(Panel **A**) Table visualization of co-occurrences of item deficits. Intense red for higher number of co-occurrences. Empty blocks represent zero co-occurrences. (Panel **B**) Graph visualization of correlation of item deficits severity. Only significant correlations (*α <* 0.05, Benjamini-Hochberg FDR correction) are visualized. Abbreviations: LOC: Level Of Consciousness; LOC-C: LOC-Commands; LOC-Q: LOC-Questions; L: left; R: right; BL: Best Language; Limb-A: Limb Ataxia, Inatt: Inattention; Facial-P: Facial Palsy. Color line thickness represent the value of Spearman’s *ρ*.

Facial Palsy and Dysarthria frequently co-occur in 68 patients. They also frequently co-occur with language deficits, with 45 patients exhibiting Facial Palsy and 34 showing Dysarthria. Both are linked with motor and sensory impairments on both sides of the body. Language deficits, specifically “Best Language”, are often associated with right-sided motor deficits, sensory issues, and visual impairments. Inattention typically accompanies left-sided motor deficits, sensory problems, and visual impairments. Level Of Consciousness-Commands (LOC-C, ability to understand and execute a simple motor command) and “Best Gaze” deficits have fewer associated with other deficits.

Figure 2, Panel **B** shows only significant correlations after Benjamini-Hochberg FDR correction for multiple comparisons (*α* = 0.05) on permutation-based p-values. Of 91 correlations, only 11 are significant after correction. The network plot reveals a strong correlation between arm and leg Motor deficits on both sides. “Best Language” shows significant correlation with LOC-Q, negative correlation with left Motor deficits (being often co-occurring with right Motor deficits). Left Motor deficits correlate significantly with Inattention, which in turn correlates with Visual deficits. Interestingly, Sensory deficits correlate significantly with Limb Ataxia.

In summary, the co-occurrence patterns and correlation network show that arm and leg Motor deficits positively covary on the same side of the body, and negatively covary on opposite sides. Language deficits may occur alone or with right Motor deficits, while Inattention covaries with left Motor deficits. Facial Palsy and Dysarthria are positively correlated and often occur with other deficits, regardless of side.

### 3.2 Repeated Spectral Clustering

Applying RSC allows to identify 5 main clusters of patients using the NIHSS data. The *k* = 5 process shows the sharpest change in the spectrum of the matrix *C*, as discussed in Appendix C. The minimum, median and maximum NIHSS profiles for the 5 clusters and the corresponding group average MRI lesion anatomy are visualized in Figure 3, Panel **B**. The Fruchterman-Reingold force-directed algorithm [33] is used for visualization so that patients who are more similar appear closer in the graph (see Panel **A**).

**Fig. 3.**
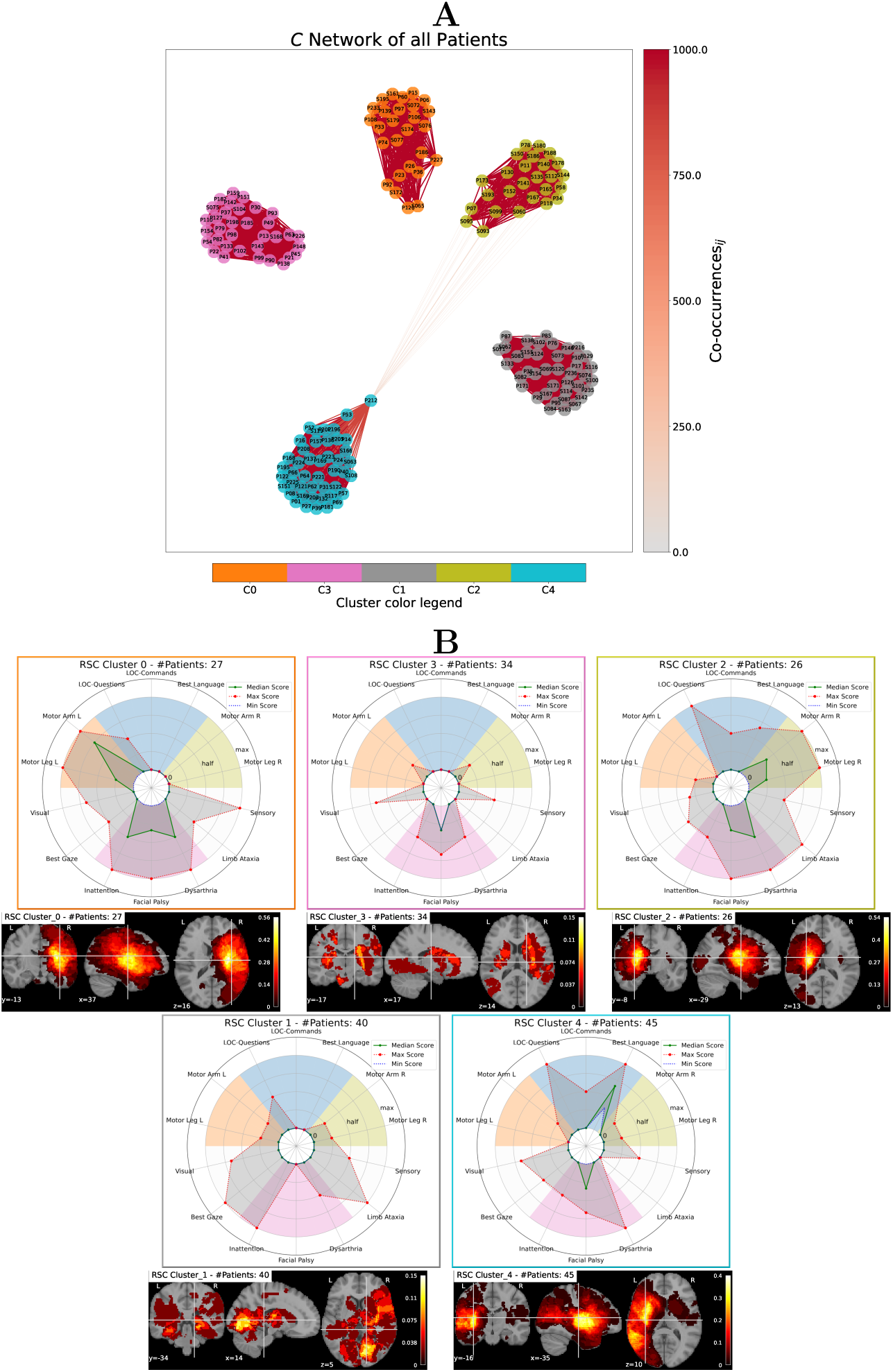
(Panel **A**) Graph visualization of the co-occurrence matrix *C*. Nodes represent patients, while colors identifies clusters. Darker red color links patients frequently clustered together. (Panel **B**) Radar charts and lesion heat maps for the clusters obtained from RSC. In the radar plots the lines represent (in blue) the minimum, (in green) the median and (in red) the maximum of the NIHSS values inside the cluster. In the heatmaps, brighter colors indicates voxels lesioned by more subjects.

Cluster 0 includes left arm-leg Motor deficits, Inattention, Facial Palsy, and Dysarthria. In the most severe cases, Visual, “Best Gaze”, and Sensory deficits can also occur (see difference between median and max deficit). The lesions localize to the vascular territory of the right middle cerebral artery (MCA).

Cluster 2 includes right arm-leg Motor deficits, Facial Palsy and Dysarthria. The lesions are roughly localized to the deep left MCA branches; in details, it involves the medial and lateral lenticulostriatal arteries of the M1 branch of the left MCA.

Cluster 4 involves selectively language deficits and Facial Palsy, and in the most severe cases can affect vision, sensation, and gaze. The lesions are localized in the territories of the superficial left MCA involving the cortical branches of the MCA, including the anterior temporal branch and the superior and inferior branches of M2.

Cluster 1 shows a high degree of deficit variability with a preference for Inattention, Visual deficits, and gaze disorders. It primarily involves the territories of the posterior cerebral artery bilaterally (both deep and superficial branches), and only partially the right MCA (bilateral posterior cerebral artery, PCA).

Finally, Cluster 3 shows a relevant presence of Facial Palsy, bilateral upper limb Motor deficits, Dysarthria and Sensory deficits. It involves the anterior choroidal arteries and penetrating branches of the MCA, Anterior Celebral Artery (ACA), and PCA bilaterally, as well as penetrating branches of the basilar artery (lacunar infarcts).

In summary, RSC provides a distribution of symptoms’ clusters confirming the presence of contralateral Motor syndromes, the relative segregation of language deficits from Motor deficits, and the association of Inattention and Sensory deficits with right hemisphere lesions.

### 3.3 Voxel-wise analysis

Applying the voxel-wise analysis allows to find statistically significant distribution of lesions coming from three clusters, in particular Cluster 0 (left Motor deficits), Cluster 2 (right Motor deficits) and Cluster 4 (language deficits). Significant regions are shown in Figure D7. The multiple-slices views are shown in the supplementary material Appendix D.

**Fig. 4.**
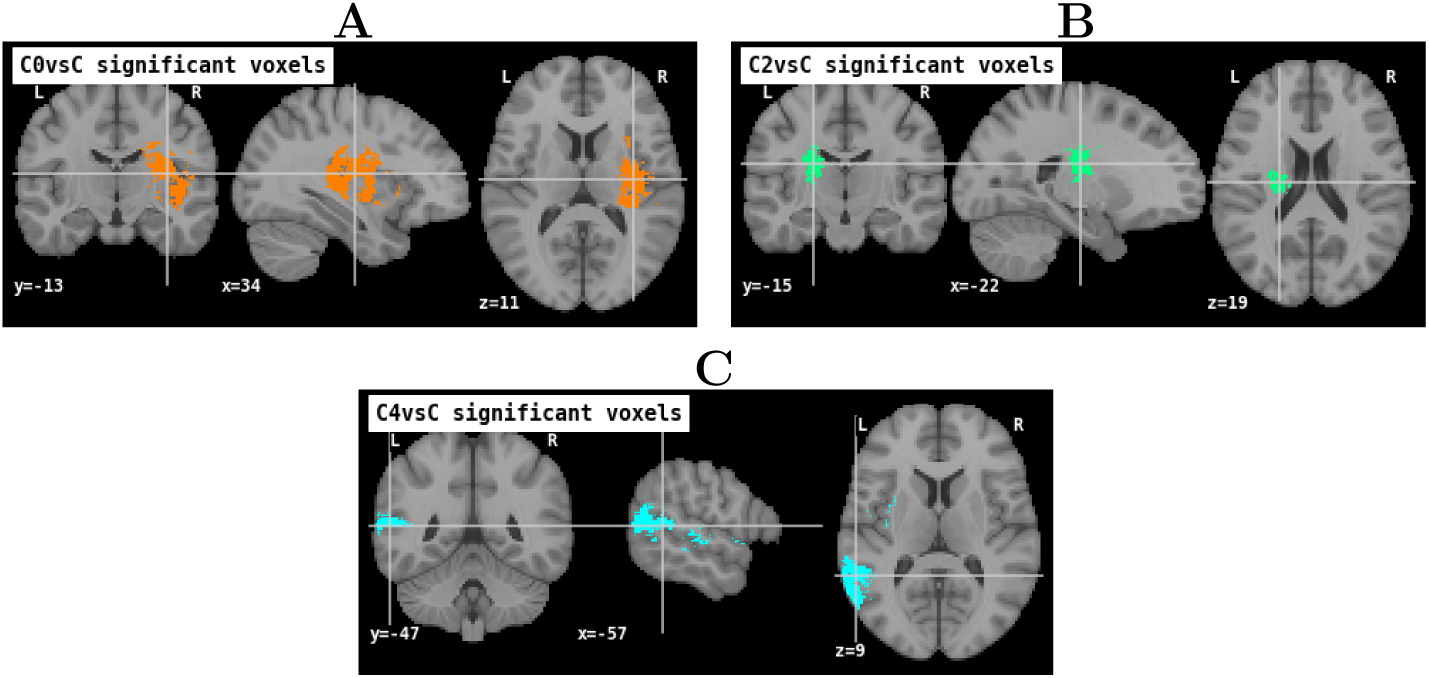
Significant voxels found from the voxel-wise analysis with FWER at *p* = 0.05. At the top (Panel **A**) in orange, Cluster 0 (left motor deficits), in the middle (Panel **B**) in green, Cluster 2 (right motor deficits) and at the bottom (Panel **C**) in light blue, Cluster 4 (language deficits).

Concerning Cluster 0, the orange region in Figure D7 Panel **A**, is part of the vascular territory of the lenticulostriate arteries of the right MCA. Anatomically, this corresponds to the right corticospinal tract at the level of the corona radiata and the posterior limb of the internal capsule, as well as the posterior insula, globus pallidus, and putamen [34].

For Cluster 2, the green region in Figure D7 Panel **B**, is part of the vascular territory of the lenticulostriate arteries of the left MCA. Anatomically, this corresponds to the left corticospinal tract at the level of the corona radiata and the posterior limb of the internal capsule.

Finally, for Cluster 4, the light blue regions in Figure D7 Panel **C**, are within the vascular territory of the inferior trunk of the left M2. Anatomically, this corresponds to the posterior insula, Heschl’s gyrus, superior temporal gyrus (Wernicke’s area), and arcuate fasciculus [35], [36].

In summary, the patients’ lesions of these three clusters are highly specific with respect to the rest of patients, and thus characteristic of these clusters.

## 4 Discussion

The clustering and imaging results are aligned with the medical literature. Interestingly, they add anatomical discriminative power to the NIHSS tests, leveraging co-occurrence statistics to distinguish similar deficits from different underlying lesions. Moreover, the application of RSC to the GDM/GSM measure, and the workflow code are themselves useful results, made available to researchers in biomedical domains.

### 4.1 The underlying structure of the NIHSS

Previous factor analyses of the NIHSS (Lyden et al. [8], Zandieh et al. [7]) identified two main factors, related to the left and right hemispheres, that can explain the majority of variance of behavioral impairment following stroke. While these results assess the conformity of the NIHSS to cerebral hemispheric lateralization, hence fundamentally validating the scale in its clinical setting, they offer limited acute stroke-phase insights for clinicians.

Here, using an unsupervised machine learning approach based on the NIHSS scores, we identify a finer-grained structure of post-stroke impairment. RSC, in addition to left and right hemispheric clusters, identifies a group of co-occurring deficits (i.e. facial palsy, bilateral upper limb motor deficits, dysarthria and sensory deficits) and anatomical correlates (i.e. bilateral basal ganglia, internal capsules) that may be associated with lacunar syndromes. This clinical entity reflects a specific etiological mechanism (i.e. arteriolosclerosis [37]) that guides the diagnostic and therapeutic algorithms for these patients. Moreover, RSC identifies numerous left hemisphere clusters, possibly due to NIHSS’s higher sensitivity to left lesions [38]. In addition, results confirm the unreliability of the limb ataxia subtest, which is scored in supra-, infra-tentorial and in bilateral lesions. This is in line with previous studies that have shown low interrater reliability of this subtest, proposing modified versions of the scale for clinical trials (see the mNIHSS [39], [40]).

Notably, RSC seems to identify a cluster (Cluster 1) with highly heterogeneous patients Figure 3. This cluster may be interpreted as patients without clear behavioral similarities (i.e. “leftovers”) according to the NIHSS, reflecting the necessity to integrate this test with additional hyperacute clinical evaluations, including cognitive and mood assessments, as shown by previous work [9], [41].

Finally, our voxel-wise analysis identifies a statistically significant set of lesions for Cluster 0 (left Motor deficits), Cluster 2 (right Motor deficits) and Cluster 4 (language deficits). In particular, motor deficits are significantly associated with corticospinal tract damage, while language deficits with damage to the arcuate fasciculus and Wernicke’s area. Interestingly, the described regions correspond precisely to anatomical substrates with a well-established prognostic role for recovery [42], [43], [44], [45], [46].

Future work, using the same validated methodological approach, could assess the longitudinal evolution and anatomical specificity of the identified clusters to better characterize post stroke recovery trajectories.

### 4.2 Unsupervised learning approaches in stroke

This work introduces a new perspective on stroke cohort data analysis. Patients are represented in a network where they are linked by similarity and clustered. This original view is partly shared by trajectory profile clustering [47] and latent class analysis[16] used in recovery studies. Statistical analyses have instead traditionally focused on variables’ correlation, variables’ covariance with imaging data, and population outcome predictions. Examples of imaging and behavioral studies can be traced to Voxel-based Lesion–Symptom Mapping (VLSM) [48] in its univariate and multivariate forms, and to the Partial Least Squares (PLS) family of applications [20]. VLSM assigns *t*-statistics, *p*-values, and correlation scores to brain regions linked with behavioral deficits, while PLS reduces and correlates dimensions of brain and behavioral data.

Overall, the traditional analyses focus on problems of the form *X^T^ X* when *X* is a matrix of *n* observations (subjects) on *m* variables; the matrix product is *X^T^ Y* when *Y* has the same *n* subjects observed, and there are *m_X_* variables in the set of features *F_X_* (e.g. behavioral scores), and *m_Y_* variables in the set of features *F_Y_* (e.g. brain voxels). In contrast, the perspective here focuses on patient profiles, patient-to-patient and patient-to-group relations, and group-specific associations of deficits and lesions, addressing the matrix product *XX^T^*. This explains the complementarity of this approach with existing literature. Data points are clustered on the set of features *F_X_* (here, NIHSS sub-scores), and cluster statistics are evaluated on the feature set *F_Y_* (binary masks of brain lesions in CT and MRI scans). The process leverages the multimodality of both *F_X_* and *F_Y_*, translating to different domains. Starting from first principles, the suitability of the GDM for ordinal data (*F_X_*) is demonstrated and used to construct the similarity network of NIHSS score profiles. The most common clustering distances include Euclidean, cosine, Manhattan, Mahalanobis and Pearson distances, as noted in [49]. Differences in pairwise similarity distributions can be found in Appendix A. Using the cosine distance would result in most patients being orthogonal, thus maximally different. However, this property is not desirable, as scoring 1 in two different items does not make two subjects maximally different, one argument being that they are both close to the healthy case where all NIHSS scores are zero. Euclidean distance is unsuitable and not theoretically motivated for the NIHSS data space, since it is a lattice space, where different distances on different axes are arguably not proportional (anisotropy and inconsistency of scale). Mahalanobis and Pearson distances are also not suitable for NIHSS data, since they assume the validity of standardization operations (mean-centering and variance-scaling) on the data, which are intrinsically skewed. Manhattan distance works well with interval data. However the GDM is still superior to Manhattan in these respects: because it accounts for the variability in each item, intensifying the impact of pairwise differences the less entropy is found in an item; because it does not depend on the magnitude of the differences, making no assumptions on the scales of different items, thus shrinking the distribution of similarities.

NIHSS items difference between two subjects conveys the difference in gravity of condition, but the gravity itself is not linear, as suggested instead by the use of single digits in the NIHSS scale. This motivates the use of GDM for such data as theoretically sound, with parsimonious assumptions, and pushing toward a more conservative position with respect to patient conditions, compared to the differences in type of symptoms. This similarity network is clustered using the unsupervised machine learning technique Repeated Spectral Clustering.

RSC unifies consensus and spectral clustering in a simple frame, taking advantage of the random initializations of *k*-means to derive robustness, as evidenced by the eigenvalue changes in *W*’s and *C*’s respective graph Laplacians. To the best of our knowledge, this is the first instance using the normalized symmetric Laplacian algorithm [27] both in the “ensemble” of random initializations constituting evidence accumulation, and as the consensus function aggregating them in a final result, requiring a single *k* choice as in classical *k*-means, supported by the spectral analysis of *W* and *C*. For related algorithms, see [50] and [28].

Future work would extend the pipeline acknowledging its limitations, enabling its application to larger stroke patients cohorts, more extensive symptom records, and other multimodal biomedical data collections, regardless of prediction targets. While the GDM’s flexibility for ordinal and continuous data is valuable, traditional preprocessing steps could support other proximity measures such as Hamming, Manhattan, and cosine distances, without hindering RSC applicability. RSC could be improved by considering geometric constraints in spectral embedding and incorporating soft partitions and probabilistic cluster attributions, currently implied by the consensus matrix. Extending these tools with a focus on interpretability can provide valuable support systems in research studies.

## 5 Conclusions

In recent years, machine learning and data science have advanced significantly, offering new prospects across many domains. The medical field’s complexity, with its numerous variables and statistical properties, makes it difficult to create data-driven mathematical models of diseases from healthcare data. In this paper, for the first time we address stroke patients clustering, presenting a novel unsupervised data analysis pipeline tailored to the properties of health scales, and applied to a leading global cause of death and disability. Many studies have analyzed patient behavior and lesion locations, but they often rely on assumptions that can limit their validity, such as treating item modalities as numerical data and using linear models. This research complements existing viewpoints by following a new and precise methodological path and introducing key elements. Our methods focus on thorough treatment of ordinal variables and avoid restrictive and distorting assumptions, using GDM and model-free lesion mapping.

These maps show anatomical separation based on cluster membership, aligned with expected localization of deficits, despite being based solely on behavioral data. They describe a detailed structure of neurological syndromes, providing additional topographical and etiological insights for clinicians in the hyperacute phase of stroke, and confirm known limitations of widely used clinical scales.

The novel and open source workflow here provided is flexible and of high methodological value, ready for extensive adaptations to multimodal biomedical data in other domains, whenever unsupervised phenotypizing of obervations is difficult but potentially enriching, and whenever clinical healthcare data comprises ordinal scales.

## Data Availability

Data can be made available upon reasonable request to Maurizio Corbettta at.

## Statements and Declarations

### Ethics approval

For data of patients of the Saint Louis cohort, this research complies with all relevant ethical regulations. Written informed consent was obtained from all participants in accordance with the Declaration of Helsinki and procedures established by the Washington University in Saint Louis Institutional Review Board. All participants were compensated for their time. All aspects of this study were approved by the Washington University School of Medicine (WUSM) Internal Review Board. For data of patients of the Padua cohort, participants from this dataset provided written informed consent following the Declaration of Helsinki principles and procedures established by the Stroke Unit and Neurological Clinic of the Hospital of Padua. This is a retrospective and non-interventional study, that received approval from the Ethics Committee of the Azienda Ospedale Università Padova.

### Availability of data and materials

The data that support the findings of this study are not openly available due to reasons of sensitivity and are available from the contributing authors upon reasonable request. Data are located in controlled access data storages at Padova Neuroscience Center and Azienda Ospedale Università Padova.

### Code availability

Code will be available upon publication at https://github.com/MedMaxLab/nihss_clustering

### Authors’ contributions

Material preparation, data collection were performed by ALB, MC, SF and LP. Data analysis was performed by MA, LFT and AZ. Medical interpretation was performed by ALB, MC and SF. Conceptualization from MA, MC, LFT and AZ. The first draft of the manuscript was written by LFT and AZ and all authors commented and edited previous versions of the manuscript. All authors read and approved the final manuscript.

## Appendix A Distances

Five different distances have been used to calculate the distributions of the pair-wise similarities, together with GDM. These five distances are the most used in clustering as discussed in [49]. Since Manhattan and Euclidean distances are not normalized in the 0-1 range, the pair-wise distance have been normalized respect the maximum distance found in the dataset.

**Fig. A1.**
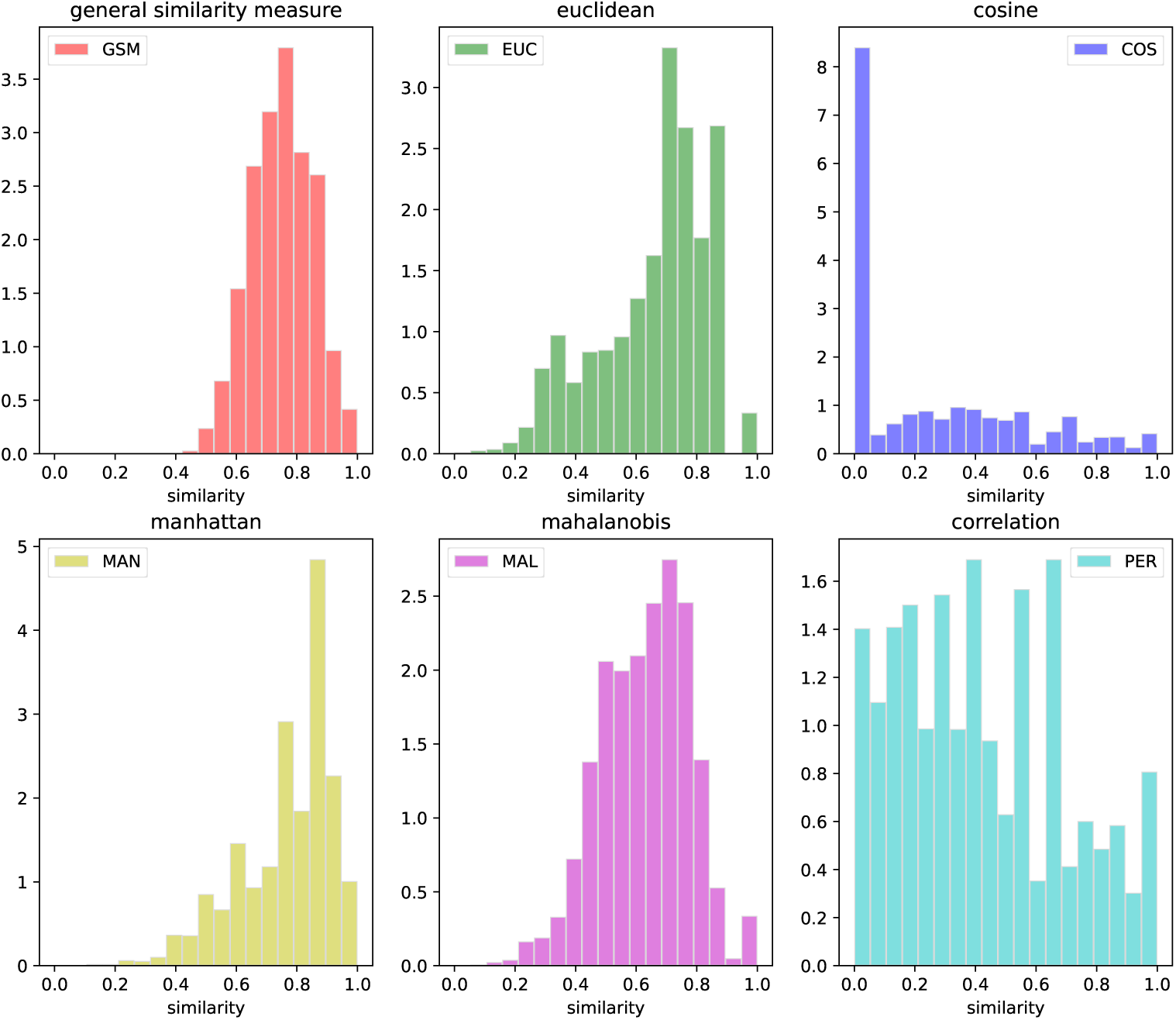
Pair-wise similarities distributions, for six different distances used. GSM stays for General Similarity Measure; EUC stays for Euclidean; MAN stays for Manhattan; MAL stays for Mahalanobis; PER stays for Pearson.

## Appendix B Other Clustering Techniques

*k*-means and hierarchical clustering are two widely used clustering techniques. This section demonstrates why these methods are unsuitable for NIHSS items and suggests alternative approaches.

Since *k* is a hyper-parameter of *k*-means, we experimented with different values ranging from 2 to 9. The raw NIHSS can be input to *k*-means, along with its normalized and standardized versions. The algorithm defaults to using Euclidean distance for this analysis. Concerning the *sklearn* ‘KMeans’ function used, the initialization was set to random, inertia to 10, and the maximum number of iterations to 300, with a random state of 12345. Table B1 reports the number of subjects for each cluster across various normalization combinations and values of *k*.

**Table B1.**
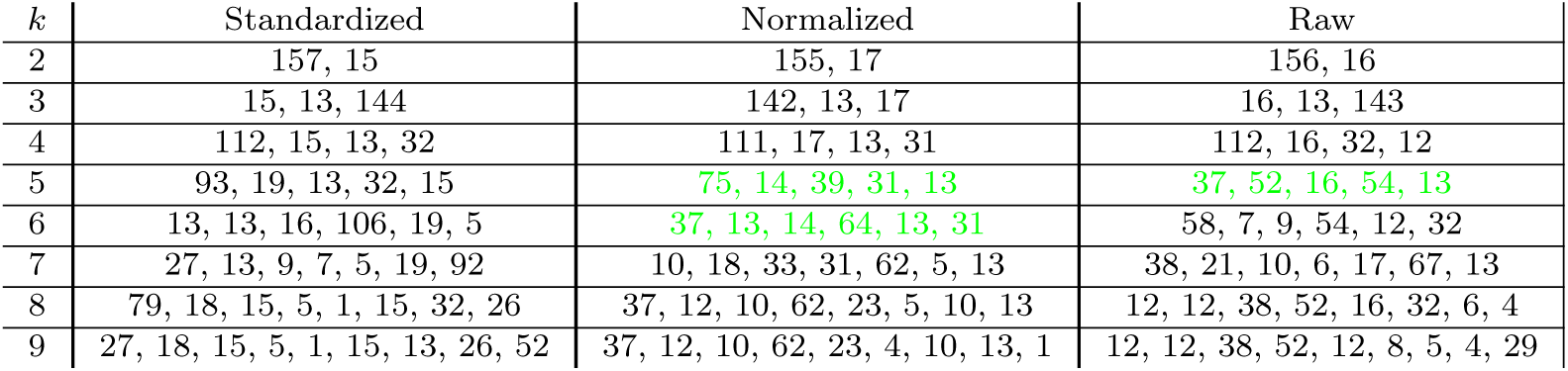
Number of subjects, for each cluster, for all the possible combinations of *k* and normalization used. In green the configurations with a number of subjects matching the 50%-5% criterion.

To evaluate cluster quality, consider cluster size: both very large clusters (over 50% of subjects) and very small clusters (under 5% of subjects) are problematic. Large clusters obscure phenotypic differences, while small clusters lack statistical power. Configurations with fewer than 8 or more than 85 subjects should be discarded. With these criteria, the only suitable configurations are normalized data with *k* = 5 and *k* = 6, and raw data with *k* = 5. In these configurations, clusters for Motor Right, Motor Left, and Language deficits were identified.

In all these three configurations the patients in Motor-R are 13, while the patients in Motor-L are respectively 14 for norm data and 16 for raw data; our clusters accounts better for patients with moderate and low scores in these items, giving 27 and 26 subjects respectively. The patients that are excluded from these three clusters, are 74 in our work, while 114 with *k*-means. The main reasons for the observed behavior could be the high-dimensionality of the data, and the spherical-shape of clusters of the *k*-means algorithm.

Hierarchical (agglomerative) clustering is another popular technique used to progressively aggregate data together, based on a distance measure. At each iteration, the subjects below a distance threshold are grouped together. The algorithm results in a dendrogram, tree-like diagram in which every data point corresponds to the leafs, and branches track the merging of subjects in larger groups. Varying the distance threshold, the tree gives different numbers of clusters from a maximum equal to the number of patients, to only one clusters of all patients. The standard technique to assess the “true” number of clusters is to check the longest interval in terms of distance threshold (GDM), for which the number of clusters does not change. The longer the interval, the stronger the evidence for a structure and number of clusters. In our case, the distance is given by the GDM matrix (thus pre-computed) and the linkage mode was set to complete. Figure B2 shows the number of clusters respect the GDM distance, where the optimal *k* was found to be 4. However following the rules before, there are the usual right/left motors and language clusters, but cluster 0 has 96 patients, that is more than half of the dataset. In conclusion, vanilla *k*-means and hierarchical clustering, are not suited for the NIHSS data of our two cohorts; spectral clustering is the next most common used clustering technique and it was used as base of this work.

**Fig. B2.**
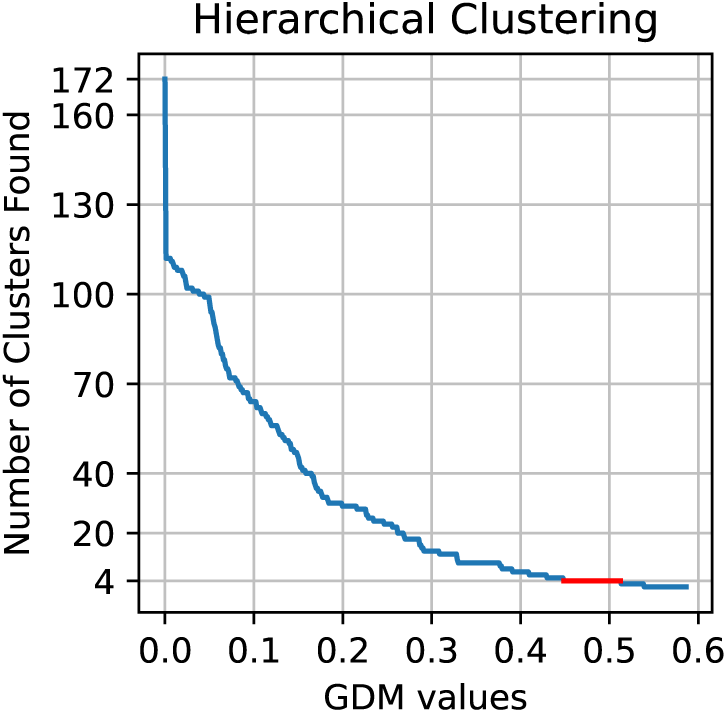
Number of clusters found by the hierarchical clustering versus the distance values. In red is shown the longest interval corresponding to 4 clusters.

## Appendix C Spectral Clustering

Spectral clustering is a very powerful clustering technique, however its geometrical foundation makes it hard to understand at first glance for non-experts in the field. In Figure C3 it is reported a brief illustrated example of how spectral clustering works.

**Fig. C3.**
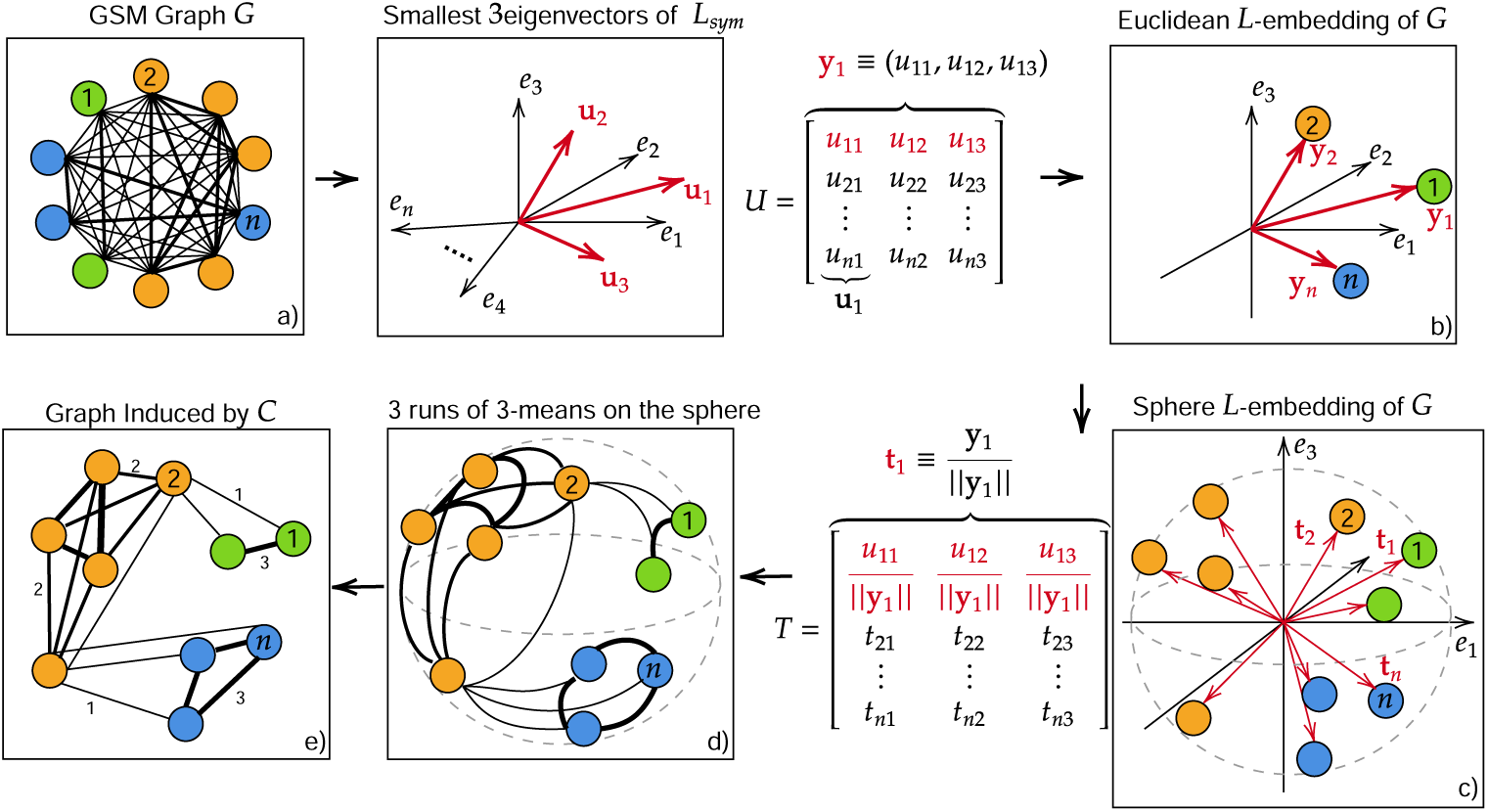
Graphical summary of how spectral clustering works in the case of *k* = 3. Starting from **a)**, the GSM matrix visualized as a graph, **b)** then the matrix *U* is computed with its respective Euclidean embedding. **c)**Then the points are projected to the unit sphere and **d)** the *k*-means is run. **e)** The *k*-means is repeated *N* times giving the final co-occurrence matrix *C* and the corresponding graph.

From the adjacency matrix *W*, built from the similarity measure with *w_ab_* = *s_ab_*, the associated normalized Laplacian matrix *L_sym_* is calculated [27]. If the graph of *W* has *n* separate sub-graphs, the Laplacian has *n* eigenvalues equal to 0. Similarly, if the Laplacian has *n* relatively small eigenvalues, the graph of *W* has *n* components far more connected internally than with one another. The eigenvectors associated to the *n* first, smallest eigenvalues are then used to create the new embedding space, where the data set is projected to. Once the spectral embedding is performed, *k*-means clustering is typically run over the new data set with *k* equal to the number *n* of eigenvectors considered. *k*-means clustering results in a complete, hard partition of the data. The above steps would complete one level of spectral clustering. However, it is well known that *k*-means results depend heavily on the random initialization of the centroids. Consequently, different runs of the algorithm will yield different results and partitions. Ensembling of clustering results, also known as consensus clustering [51], is the process of aggregating different partitions from multiple algorithms into a single, robust one. Evidence Accumulation Clustering [29] is a particular consensus clustering technique. In particular, this technique is based on repeating *N* times the *k*-means algorithm. Through repetitions, evidence accumulates regarding which data samples consistently and reliably cluster together. The so called co-occurrence matrix *C* is defined, where the entries *C_ab_* are the number of times two subjects *a* and *b* are clustered together over the *N* trials, as shown in Figure C4.

**Fig. C4.**
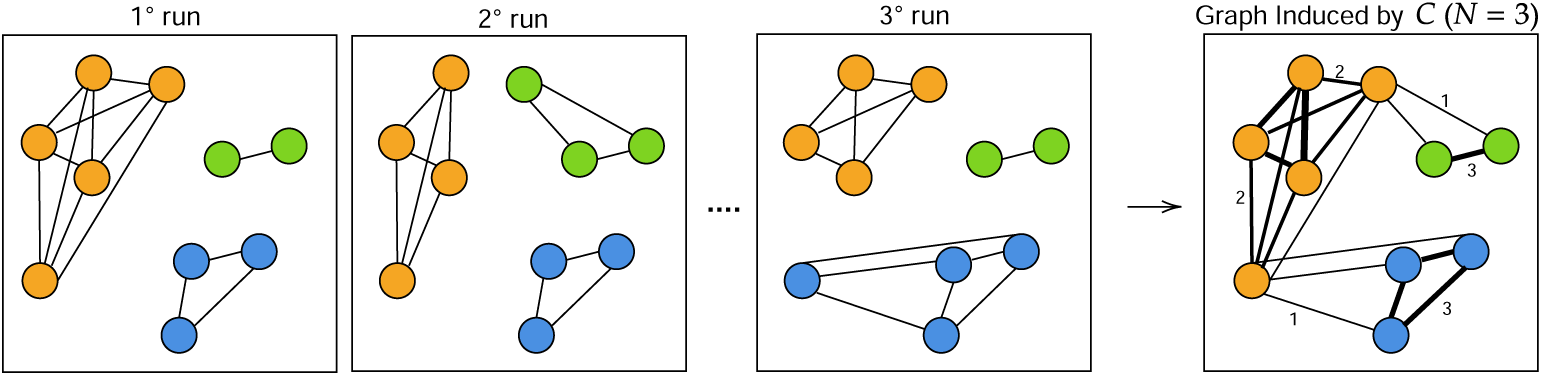
Toy example of how the *C* matrix is built. Here different data points are clustered together differently for each run of the *k*-means. At the end the *C* matrix is obtained by counting the number of times a link is present between two nodes over *N* runs.

The matrix *C* describes a new graph, where strong connections represent frequent associations in the same clusters. The default hard partition from *k*-means is lost to a soft partition of connected communities. Since the objective of clustering is still assigning labels to data points, a further clustering method could be applied to the co-occurrence matrix *C*, to finally separate the communities from the new co-occurrence network. In this work a further spectral embedding and *k*-means is performed on *C*, given that its spectral properties and separability are enhanced compared to the original adjacency matrix *W*. This idea is analogous to work in [28], where the spectral embedding relies on a different Laplacian form, *L_rw_*, and the clustering for evidence accumulations are several and different algorithms. The way in which *n* is chosen for *W* and *C* relies on the spectrum of the normalized Laplacian of the matrix *C*. As stated above, *n* null eigenvalues indicate *n* completely separate groups in the data graph, and *n* very small eigenvalues followed by a larger eigenvalue indicate *n* softly distinct groups. So, in the absence of exactly 0-valued eigenvalues, the problem consists in finding a spectral gap, i.e. a large difference between an eigenvalue and all the other quasi-0 predecessors. The spectral-gap is measured as the difference between the *k*-th and the *k* + 1-th ordered eigenvalues of the Laplacian matrix of *C*. At different values of *n*, *k* when working on *W*, one can end up with different spectral gaps in the spectrum of *C*. In Figure C5 is reported the spectral-gap profile for different *n*, *k* trials. As can be seen, for *k* = 5 there is the highest values of the spectral gap: by the max-gap criterion it is the suggested choice.

**Fig. C5.**
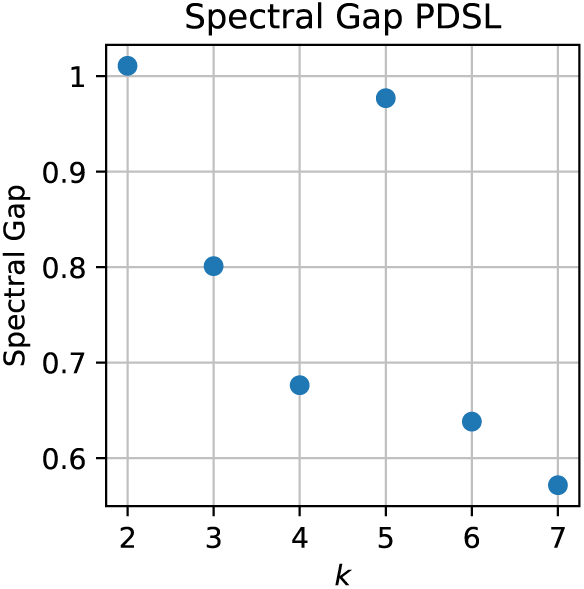
Spectral-gap profile for various *k*, for the Padua and St. Louis dataset together.

### C.1 Statistics

Considering the number of times *N* the *k*-means has to be executed, the event *two patients are clustered together* is a dichotomous event; hence *C_ab_* follows a Binomial distribution. Since *C* can be written as *C* = *NP, C_ab_* are interpreted as mean values, so the error associated to each entry scales as 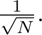 From statistical reasoning on the error of the variance, the minimum number of trials is therefore 200; in this work *N* = 1000. Going into the details of the *k*-means function implemented in *scikitlearn*, there is a parameter called *n_init_* that indicated how many runs have to be executed to give the final answer, i.e. the labels. The idea behind is to try different runs and choose that with smallest inertia, as best candidate out of the batch. Further details can be seen in the corresponding web page https://scikit-learn.org/stable/. Since in this work the *k*-means is repeated several times in order to fully exploit the stochasticity of the centroids initialization, in principle *n_init_* = 1. However as can be seen in Figure C6, moving from *n_init_* = 4 to *n_init_* = 1 gives rise to a bell-like shape probability distribution function, centered in 54; still the main peak of the inertia is in the bins [42, 43]. This indicates that increasing *n_init_* a little bit (e.g. 4) is beneficial for having clusters with low inertia and avoid to have noisy-configurations with inertia around 54. However even in the case with *n_init_* = 1 the final labels are equal to the case of *n_init_* = 4.

**Fig. C6.**
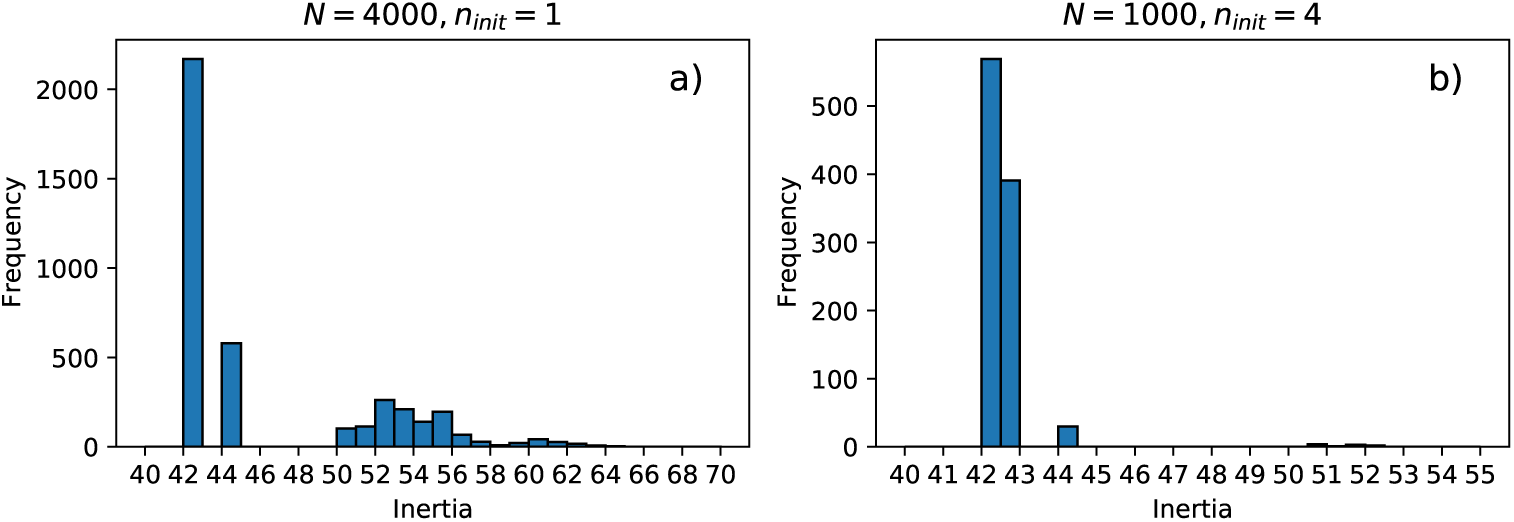
Comparison between histograms of the inertia in the case *n_init_* = 1 a) and *n_init_* = 4 b). The total number of runs is thus fixed to *N* ∗ *n_init_* = 4000.

## Appendix D Voxel-wise Results

The statistical significant lesions shown in Figure D7, are here visualized at different slices.

**Fig. D7.**
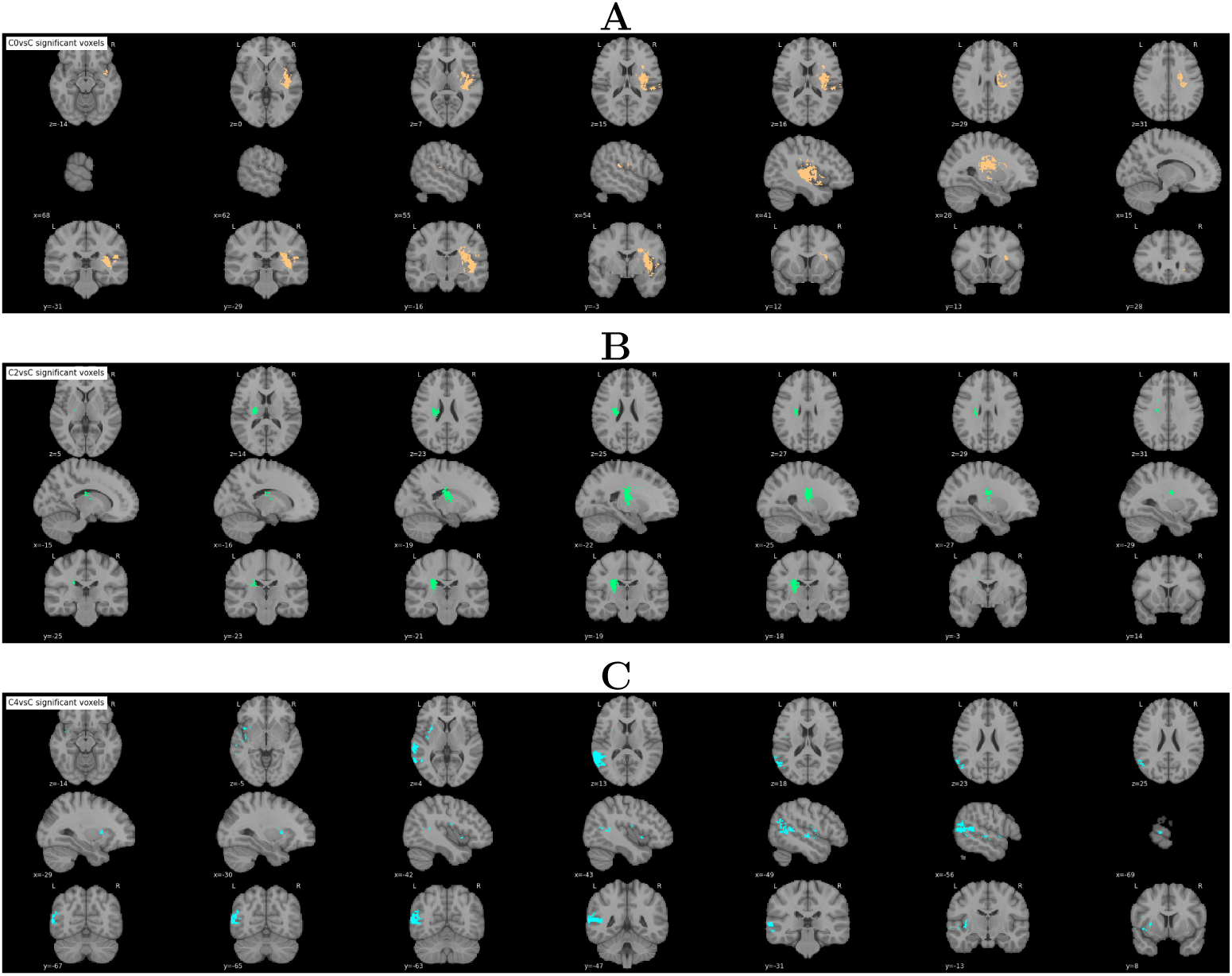
Significant voxels found from the voxel-wise analysis with FWER at *p* = 0.05 for different slices. At the top (Panel **A**) in orange, Cluster 0 (left motor deficits), in the middle (Panel **B**) in green, Cluster 2 (right motor deficits) and at the bottom (Panel **C**) in light blue, Cluster 4 (language deficits).

